# Polydrug overdose mortality caused by synthetic opioids and stimulants: Current sex- and age- specific trajectories in United States national data for 2018-2024

**DOI:** 10.1101/2025.08.21.25334165

**Authors:** Eduardo R. Butelman, Yuefeng Huang, Siri Shastry, Alex F. Manini, Rita Z. Goldstein, Nelly Alia-Klein

**Author notes:** **Corresponding Author:** Dr. Eduardo Butelman Neuropsychoimaging of Addictions and Related Conditions, Department of Psychiatry, Icahn School of Medicine at Mount Sinai, 1 Gustave L. Levy Place, New York, NY 10029-5674 **E-mail:**.

## Abstract

Recent years have shown increases in overdose (OD) mortality caused by *polydrug* exposure to synthetic opioids such as fentanyl and stimulants such as methamphetamine or cocaine. The goal of this study is to understand the recent trajectory in this polydrug OD mortality, especially associated with decedents’ sex and age. We carried out a cross-sectional analysis of national data for persons aged 15-74, from CDC WONDER for 2018-2024 (data for 2024 are considered provisional at the time of analysis). The outcome measure was OD mortality/100,000 population; annual data for 2018-2024 were analyzed with joinpoint regression, and the most recent years (2023-2024) were analyzed with ANOVA and multiple linear regression. Males had greater polydrug OD mortality compared to females, across 2018-2024. Sex-specific joinpoint regressions detected increases in polydrug OD after 2018, then decreases from 2023 in males, and from 2022 in females. For these polydrug OD, the mean annual percent change (APC) in 2024 versus 2023 was -36% and -31% in males and females, respectively. For synthetic opioids *without* stimulants, OD trends in 2024 versus 2023 were similar to those for polydrug OD (-42% and -39% APC in males and females, respectively). However, OD for stimulants *without* synthetic opioids showed relatively smaller changes (-3% and -2% APC in males and females, respectively). Stratification into 10-year age groups for polydrug OD revealed that mortality peaked at age 35-44 and then declined at older ages. Recent decreases in polydrug OD mortality were observed across age groups, with joinpoints detected in 2022 or 2023. These findings indicate that after increases from 2018 onward, polydrug OD mortality caused by synthetic opioids and stimulants exhibited substantial decreases in both males and females in the most recent data for 2024, across a broad age range. Because relatively small changes were observed in OD mortality caused by stimulants *without* synthetic opioids in this time period, the decreases in polydrug OD mortality are more likely to be caused by changes in exposure, prevention or intervention strategies focused on opioids rather than on stimulants. While this polydrug OD mortality has decreased in 2024, it remains at concerning levels.

## Introduction

The United States has undergone an epidemic of overdose (OD) mortality in recent years, exacerbated during the COVID pandemic[1–3]. These high OD rates are caused primarily by synthetic opioids such as the mu-opioid receptor (MOR) agonist fentanyl or its analogs [4,5], and by stimulants such as methamphetamine or cocaine[6–9]. Importantly, the current “fourth wave” of OD has shown increases in *polydrug* exposure to these two types of compounds[7,10,11]. For example, there was a 37% increase in the annual rate of nonfatal polydrug OD caused by opioids and amphetamines, for 2019 versus 2018[12]. In addition to the risk of *knowingly* using both opioids and stimulants [13,14], there has also been a concerning increase in OD due to the concealed addition of synthetic opioids such as fentanyl in the drug supply, including counterfeit oral formulations[15,16], as well as into other illicit formulations, which are then injected, inhaled or smoked[17].This concealed presence of synthetic opioids can further increase the risk of OD morbidity and mortality[18–20].

Overdose morbidity and mortality caused by MOR agonists is principally due to respiratory depression[21,22], whereas those caused by stimulants (mediated principally through inhibition of monoamine neurotransmitter reuptake)[23,24] are due to different mechanisms, such as cardiovascular, neurovascular or neurological insults[25,26]. Opioid OD are acutely managed by administration of MOR antagonists, principally naloxone. Notably, naloxone has become more widely available in 2023-2024, due to the approval of several over-the-counter intranasal formulations, expanding accessibility in the community, compared to prior years[27,28]. Furthermore, receipt of medications for opioid use disorder (MOUD, such as buprenorphine or methadone) also provides increased safety and decreased OD events, although MOUD receipt and adherence can vary based on social determinants of health[29–31]. In recent years there has also been increased emphasis on “low threshold” referrals for MOUD[30,32], as well as initiation of this treatment in emergency settings[33]. By contrast, there are no medical countermeasures or antidotes specifically approved for OD caused by methamphetamine or cocaine [34]. Some preclinical studies have delineated acute pharmacological interactions between synthetic opioids and stimulants (e.g., on respiratory depression)[35]. While the pathophysiological mechanisms underlying morbidity and mortality in polydrug OD are not fully understood at the clinical level [10,36,37], it is possible that the risk of cardiac arrhythmia could be prominently increased after dual exposure due to the concomitant respiratory depression and hypoxia caused by MOR agonists [22,38] and the increased myocardial oxygen demand caused by stimulants [39,40].

Based on the state of the field, the main aim of this study is to determine the latest trajectory of OD caused by polydrug synthetic opioid and stimulant exposure, as this has not been directly examined in recent studies[41–43], based on national data from the United States Centers for Disease Control and Prevention (CDC). A second aim of the study is to determine how the changes in polydrug OD mortality occurred based on major individual differences in variables such as sex and age, which we and others have shown to be highly relevant to this risk[6,41].

## Methods

This study used de-identified publicly available data on “multiple cause mortality”, from the CDC WONDER platform for 2018-2024 (https://wonder.cdc.gov/mcd.html). The Institutional Review Board of the Icahn School of Medicine at Mount Sinai certified that this study is not considered Human Subjects Research, and is thus exempt from its review requirements. As part of confidentiality protections, data that were “suppressed” (due to nl<l10 per cell) or deemed “unreliable” in CDC WONDER were excluded from analysis. Data for 2024 are considered provisional at the time of downloading (June 2025). The overall age range studied was 15-74, due to low prevalence at younger and older ages. This observational study followed STROBE reporting guidelines (see supplement). The overall level of significance, alpha, was set at the p=0.05 level.

### Data set

The main outcome variable was age-adjusted death rate per 100,000 population. We examined ICD-10 codes for overdoses (unintentional X40-X44, suicide X60-X64, homicide X85, and undetermined intents Y10–Y14). Data were obtained for three drug categories: “other synthetic narcotics” (“synthetic opioids hereafter; this predominantly reflects fentanyl and its analogs, but also compounds from other chemical scaffolds; T40.4)[21,44], psychostimulants with abuse potential (which predominantly reflects methamphetamine but also includes its analogs; T43.6), and cocaine (T40.5). Data were stratified by sex for the main analyses. In follow-up comparisons we also examined the two largest racial groups (White and African American or Black) in stratified analyses for synthetic opioid and stimulant polydrug overdoses. A further follow-up also stratified six consecutive 10-year age groups (i.e., 15–24, 25–34, 35–44, 45–54, 55–64 and 65–74). When this age stratification was carried out, analyses were based on crude rates.

### Data analyses

Annual data were obtained initially for polydrug OD mortality caused by synthetic opioids and stimulants, by searching: “Other synthetic narcotics” (i.e., synthetic opioids, ICD-10 code T40.4) *AND* [“Psychostimulants with abuse potential” (T43.6), *OR* cocaine (T40.5)]; this latter composite category is termed “stimulants” hereafter, for brevity. The CDC WONDER platform does not have a “*NOT”* search function, therefore in order to examine OD that were caused by synthetic opioids *without* stimulants (and vice versa), we carried out the following re-calculations, based on a simple Boolean strategy:

***Group a) Synthetic opioids without stimulants*** *= [Synthetic opioid] - [Polydrug Synthetic opioid + Stimulant]*

***Group b) Stimulants without synthetic opioids*** *= [Stimulant] - [Polydrug Synthetic opioid + Stimulant]*

Whenever relevant, standard errors (SE) for rates/100,000 for Groups a) and b) above were calculated with a Poisson approximation: SE= 100,000 / N_Population_ * √[OD counts_Group a_ + OD counts_Group b_].

### Joinpoint Analysis

Joinpoint Regression software (V 5.4.0)[45] from the National Cancer Institute was used to examine annual trends in log-transformed age-adjusted overdose mortality rates (per 100,000). The technique was used to determine trends across years in a data-driven manner, by detecting “joinpoints” when a significant change occurs. Jointpoint regression examines annual percent change (APC) in the rate of OD for consecutive years (APC= ((Rate_Year N_ - Rate_Year N-1_)/Rate_Year N-1_) X100%). Thus, a positive APC value indicates an increase in OD mortality over consecutive years, and a negative APC indicates a decrease. The joinpoint regression parameters were as follows: We set the maximum number of possible joinpoints=2, taking into consideration the number of years under study[46]. The first order autocorrelation option was used to account for dependence of time series data. The Bayesian Information Criterion was used to select the number of joinpoints (i.e., 0-2) in the optimal model. The empirical quantile method[47] was used to calculate 95%CI for the APC, with 5001 resamples and a seed=10000.

### Follow-ups and sub-group analyses

#### a) Focus on the most recent data

OD mortality rates in 2024, and annual percent changes in OD mortality in 2024 versus 2023 were initially examined, with 2-way ANOVA (sex X type of drug), using US census division - level data, after examining normality with the D’Agostino-Pearson test and checking quantile-quantile plots (Graphpad Prism V10.5.0). These ANOVAs used the Greenhouse-Geisser correction for violations of sphericity. In order to examine if major sociodemographic variables and public health interventions[48] were associated with state-level annual percent change in polydrug OD mortality rates for 2023-2024, we carried out an ordinary-least squares multiple linear regression, with asymmetric profile-likelihood 95%CI for the parameters. The predictor variables were sex (with females as the reference category), and z-score transformed values for prescribed naloxone dispensing rate per 100 persons (2023 CDC data)[49], and the following major social determinants of health, based on state-level data from the US Census Bureau’s American Community Survey from 2023[50]: median household income based on census, OD rates in 2023,%of population which is African-American or Black, and % of population without health insurance. Utility of simpler regression models was evaluated with the Akaike Information Criterion. In this analysis, data from the following states were excluded, due to data suppression or unreliable estimates: Hawaii, Montana, Nebraska, New Hampshire, North Dakota, South Dakota, Vermont and Wyoming.

#### b) Stratification by age

In a followup examining national data in 10-year age groups (overall age range 15-75), the above joinpoint procedure was also used for 2018-2024, examining crude rates.

#### c) Stratification by race

We carried out stratified joinpoint analyses as above, for the two largest racial groups in national data (African-American/Black and White).

## Results

### Demographics and mortality rates

Supplementary Table S1A-C shows the number of OD deaths caused by the drug types under study, together with crude rates and age-adjusted rates, stratified by sex and year. Overall, males had approximately 2-fold greater overdose rates compared to females across the three drug types causing the OD, and years under study.

### Joinpoint regressions of national data

We first examined sex-specific trajectories with joinpoint regressions (Fig. 1 and Table 1), which yielded differences across drug type and sex.

**Figure 1:**
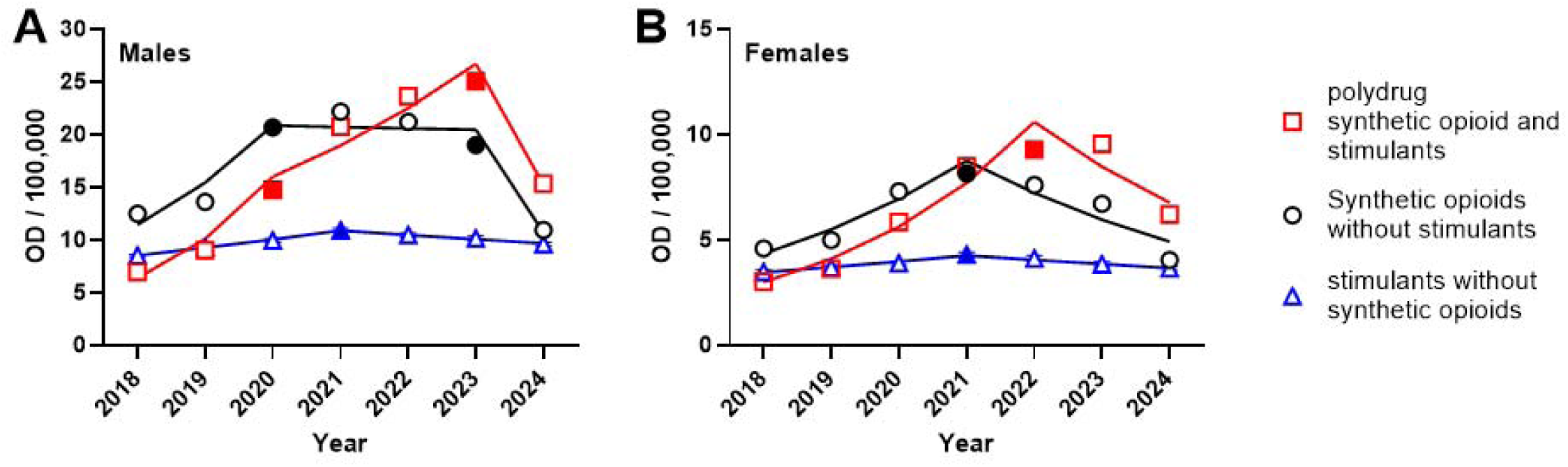
Age-adjusted overdose (OD) mortality rates / 100,000 census population for 2018-2024 for males and females (A-B, respectively). *Note different Y-axis ranges in the two panels.* Regression lines are based on optimal joinpoint regression results (see Table 2). Standard error (SE) bars typically fall within the symbols. Filled symbols indicate the “joinpoints” for each type of overdose based on the optimal models (i.e., the year in which a change in trend is observed). Full data are in Supplement Table S1A-C.

**Table 1:** Joinpoint regression optimal models in annual overdose mortality rates (per 100,000; age adjusted). Models illustrated in Fig. 2.

| **1A: Polydrug: Synthetic opioids *and* stimulants** |  |  |  |  |  |  |  |  |  |
| --- | --- | --- | --- | --- | --- | --- | --- | --- | --- |
|  | Trend 1 |  |  | Trend 2 |  |  | Trend 3 |  |  |
|  | Years | APC*<br>(95%CI) | P-value | Years | APC<br>(95%CI) | P-value | Years | APC<br>(95%CI) | P-value |
| **Males** | 2018-2020 | 58.6<br>(17.7 - 305.0) | p= 0.013 | 2020-2023 | 18.7<br>(9.3 - 43.3) | p=0.008; | 2023-2024 | -43.1<br>(-69.3 - -16.7) | p=0.0004; |
| **Females** | 2018-2022 | 37.2<br>(23.5 - 88.2) | p< 0.000001 | 2022-2024 | -20.0<br>(-63.8 - 2.5) | NS | N/A: only 2 trends in optimal model |  |  |

\*APC: Annual percent change; positive values indicate increases in year-to-year in OD mortality, negative values indicate a decrease; see Methods

| **1B: Synthetic opioids *without* stimulants** |  |  |  |  |  |  |  |  |  |
| --- | --- | --- | --- | --- | --- | --- | --- | --- | --- |
|  | Trend 1 |  |  | Trend 2 |  |  | Trend 3 |  |  |
|  | Years | APC*<br>(95%CI) | P-value | Years | APC<br>(95%CI) | P-value | Years | APC<br>(95%CI) | P-value |
| **Males** | 2018-2020 | 35.0<br>(23.3 - 90.8) | p< 0.000001 | 2020-2023 | -0.6<br>(-8.1 - 13.0) | NS; | 2023-2024 | -47.7<br>(-66.1 - -27.4) | p< 0.000001; |
| **Females** | 2018-2021 | 26.3<br>(14.4 - 46.9), | p< 0.000001; | 2021-2024 | -17.6<br>(-33.0 - -8.3) | p=0.0004; | N/A: only 2 trends in optimal model |  |  |

| 1C: Stimulants *without* synthetic opioids |  |  |  |  |  |  |  |  |  |
| --- | --- | --- | --- | --- | --- | --- | --- | --- | --- |
|  | Trend 1 |  |  | Trend 2 |  |  | Trend 3 |  |  |
|  | Years | APC*<br>(95%CI) | P-value | Years | APC<br>(95%CI) | P-value | Years | APC<br>(95%CI) | P-value |
| **Males** | 2018-2021 | 8.5<br>(8.1 - 8.9) | p<<br>0.000001; | 2021-2024 | -3.9<br>(-4.3 - -3.6) | p<<br>0.000001; | N/A: only 2 trends in optimal model |  |  |
| **Females** | 2018-2021 | 7.3<br>(4.1 - 12.8) | p<<br>0.000001; | 2021-2024 | -4.9<br>(-12.7 - -1.8) | p=0.0028; | N/A: only 2 trends in optimal model |  |  |

**Table 2:** Multiple linear regression for annual percent change in mortality for polydrug OD from 2024 versus 2023, studying sex and state-level predictors.

| Variable | Beta Estimate | 95% CI<br>(profile likelihood) | P value |
| --- | --- | --- | --- |
| Intercept | -31.72 | -35.29 to -28.14 | <0.0001 |
| Sex<br>(Male; Female as<br>reference) | -8.405 | -17.67 to 0.86 | 0.075; NS |
| State-level naloxone<br>dispensing rate/100<br>(2023) | -1.16 | -3.79 to 1.47 | 0.38; NS |
| State-level median<br>household income (\$) | 5.29 | 2.54 to 8.04 | 0.0003 |
| State-level % without<br>health insurance<br>coverage | 6.30 | 3.44 to 9.15 | <0.0001 |
| State-level %Population<br>that is African-American /<br>Black | -4.84 | -7.38 to -2.28 | 0.0003 |
| State-level OD<br>rate/100,000 (2024) | 0.12 | -0.15 to 0.38 | 0.39; NS |

### Polydrug OD caused by synthetic opioids *and* stimulants

In males, there was an initial increase in OD from 2018, with two joinpoints (2020 and 2023), followed by a decrease in 2024. In females, there was also an increase from 2018, with a single joinpoint in 2022. **Synthetic opioids *without* stimulants:** In males, there was an initial increase in OD from 2018, with two joinpoints (2020 and 2023), showing a period of relative stability (possible plateau), followed by a decrease in 2024. In females, there was also an increase from 2018, with a single joinpoint in 2021, and a decrease thereafter. **Stimulants *without* synthetic opioids:** For both males and females, there was only one joinpoint (2021), indicating an increase in OD mortality in 2018-2021, followed by a decrease in 2021-2024. Of note, the magnitude of the APC was smaller for stimulants *without* synthetic opioids, compared to the two other drug types, indicating relative stability across 2018-2024 of OD caused by stimulants without synthetic opioids, compared to the former two groups (see Fig. 1).

### Overdoses mortality in the most recent year, 2024

We followed up the above analyses with a focus on the last year available (2024; provisional data), using census division - level data (full data with individual census division in Table S2A-C). A 2-way ANOVA for 2024 (Figure 2A) showed main effects of sex and of drug type, and no significant sex X drug interaction. Tukey tests show that polydrug OD rates in 2024 were greater than those caused by the two other categories (which did not differ from each other). This finding held in both males and females.

**Figure 2:**
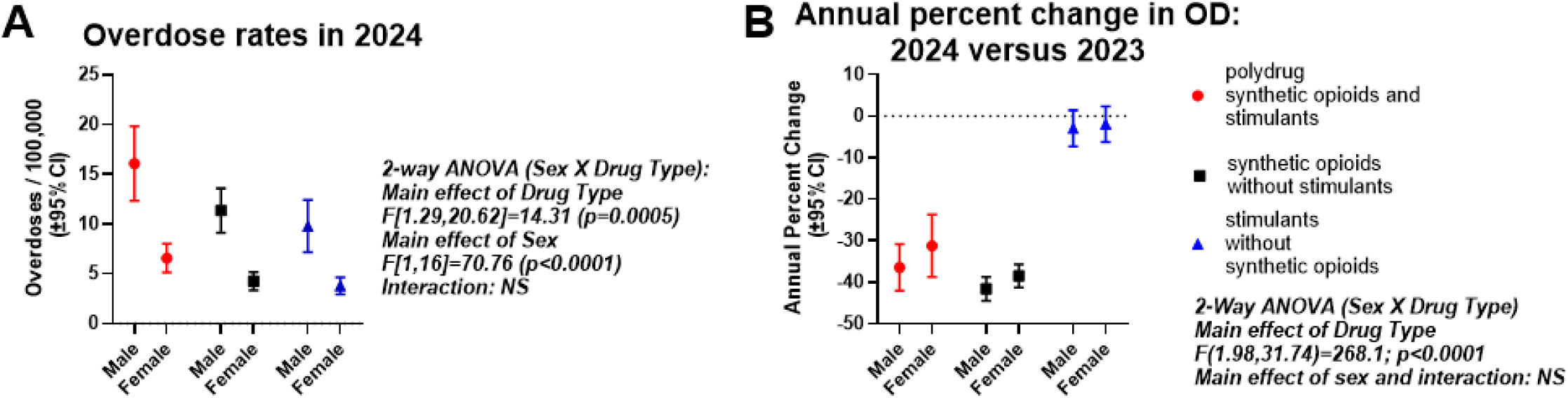
**A.** Age-adjusted overdose mortality rates in the most recent year (2024) in males and females, based on census division - level data. **B.** Annual percent change (APC) in 2024 versus 2023. Full data in Table S2A-C

### Examination of APC data for 2024 versus 2023

There were greater decreases in OD mortality in the polydrug and synthetic opioid without stimulant groups, compared to the stimulant without synthetic opioid group (Fig. 2B). Thus, a 2-way ANOVA using census division-level data detected a significant main effect of drug type, but no effect of sex or sex X drug interaction (Fig. 2B). Post hoc Sidak tests show that there was a greater decrease in OD rates in 2024 vs 2023 for the polydrug OD, and synthetic opioid without stimulant groups, versus the stimulant without synthetic opioid group, both in males and females.

### Multivariable adjusted model for annual percent change in polydrug overdoses in 2024 versus 2023

We then extracted state-level OD data (Table S3) and sociodemographic data (Table S4), and carried out a univariate Spearman correlation matrix (Table S5) for an initial univariate exploration. This was followed by a multiple linear regression, with state-level APC data for 2024 versus 2023 as the outcome (e.g., Fig 2B), with a multiple linear regression, adjusting for sociodemographic variables. These predictor variables explained a substantial proportion of the variance in the outcome variable (R^2^=0.36; df=79). As shown in Table 2, neither sex, state-level naloxone dispensing rates and OD rate at baseline (2023) were not significant predictors of APC. The adjusted p-value for sex was marginal (p=0.075), and in a follow-up simple linear regression, we again found a marginal effect of sex on this APC (p=0.1; not shown). However, greater household income, and greater %of persons without health insurance were positive predictors of APC (indicating lesser decreases in OD rates). Greater %African-American or Black population was a negative predictor of APC (indicating greater decreases in OD rates). Simpler models (e.g., without median household income), were less preferred, based on the Akaike Information Criterion (not shown).

### Age stratification of polydrug OD mortality in 2018-2024

We stratified crude mortality data in consecutive 10-year age groups (overall age range 15-74; Fig. 3), and carried out separate joinpoint analyses in males and females. There was a biphasic pattern of age-related mortality across the years, with relatively low levels in the youngest age group (15-24), increasing until age 35-44, and then declining gradually from 45-54 to 65-74; this overall pattern was observed both in males and females. Sex differences (OD mortality M>F) were generally observed at each age group, and across years. The number and years of joinpoints differed across males versus females, indicating sex differences in the trajectories of these polydrug OD. Full data and jointpoint regression results are in Table S6.

**Figure 3:**
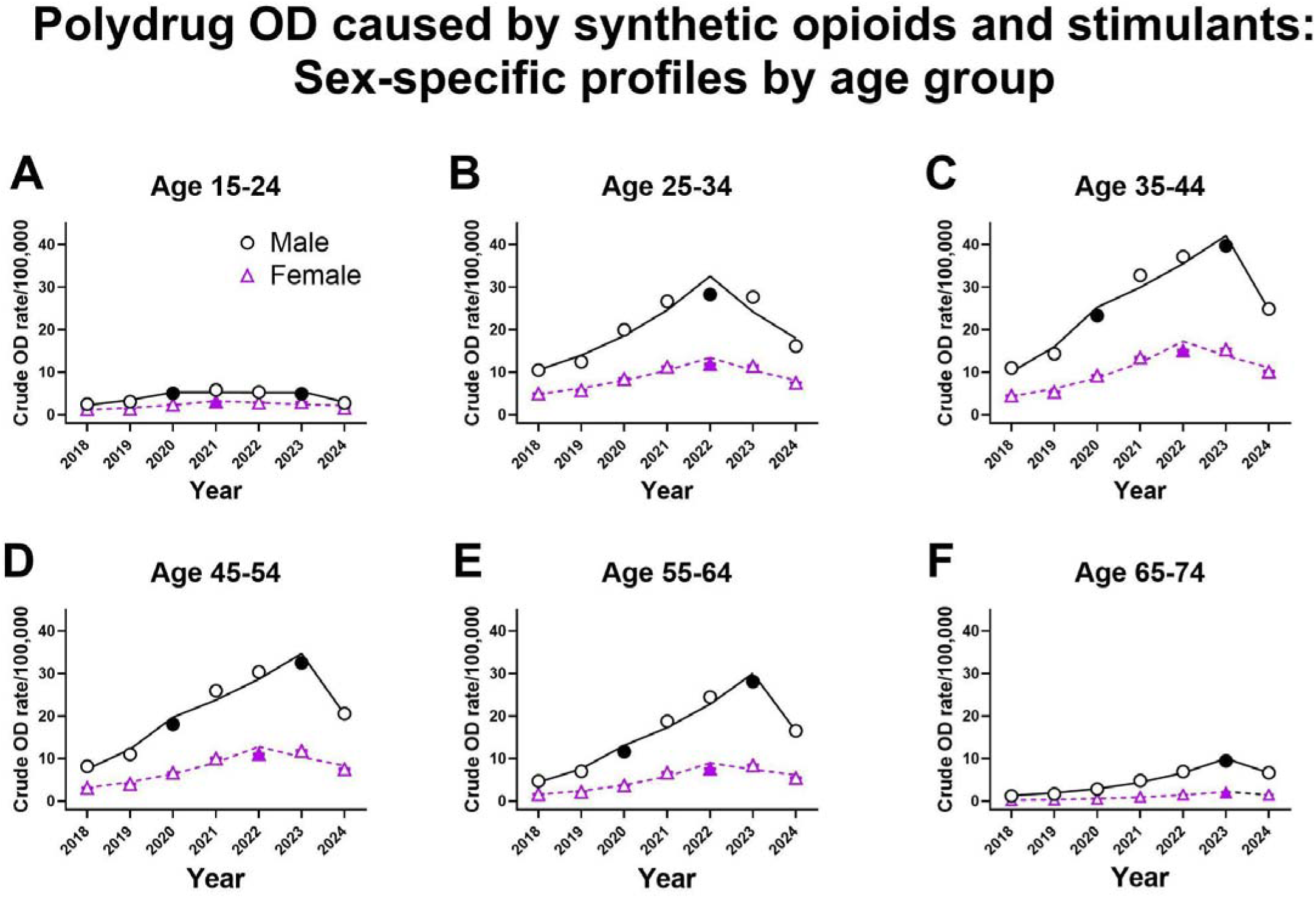
Crude OD mortality rates / 100,000 for 2018-2024 for males and females across 10-year age groups, (A-F). Some standard error bars fall within the symbols. Lines are based on optimal joinpoint regression results; filled symbols indicate the optimal “joinpoints”. Full data are shown in Table S6.

### Race stratification of polydrug OD mortality in 2018-2024

We carried out sex-stratified joinpoint analyses for the two most prevalent race categories in national data (African-American/Black and White). Peak mortality occurred in 2022-2023. While there were greater peak OD rates of persons in the African-American/Black category versus white, the APC in 2024 versus 2023 were similar. For example, in males there was a calculated APC of -49% [95%CI: -20% - -71%] for persons in the African-American/Black category, and -42% [95%CI: -20% - -56%] in the white category (data not shown).

## Discussion

Polydrug OD caused by synthetic opioids and stimulants have reached epidemic proportions in recent years[7,51]. In this national cross-sectional study, joinpoint regression was used to examine the trajectory of this polydrug OD mortality across 2018-2024, expanding from prior reports [43,52]. This analysis detected increases in polydrug OD mortality after 2018, until 2023 in males and 2022 in females, and declined thereafter. Focusing more directly on the most recent trends (i.e., 2024 versus 2023; Figure 2B; Table S2A), relatively large decreases were observed in this polydrug mortality (i.e., -36% and -31% APC in males and females, respectively). Similar decreases were detected in 2024 versus 2023 for OD caused by synthetic opioids *without* stimulants (i.e., -42% and -39% APC in males and females, respectively), whereas smaller changes were observed for OD caused by stimulants without synthetic opioids (i.e., -3% and -2% APC in males and females, respectively). This result suggests that recent decreases in OD mortality for these types of compounds may be due primarily to changes in exposure, and/or public health prevention and intervention strategies focusing on synthetic opioids [30,53,54]. For example, changes in the illicit market supply for synthetic opioids have been observed (e.g., dosage or purity of fentanyl) [16,55]. Enhanced prevention and intervention strategies also included information campaigns, broader availability of intranasal naloxone [56,57], expansion of availability of fentanyl test strips [58], community-based drug checking services [59,60], as well as increased emphasis on rapid initiation and/or referral for MOUD in emergency settings [61–63], and reduction in regulatory barriers to MOUD prescribing [30,33]. Different cohort studies show that MOUD (e.g., methadone or buprenorphine) results in significant protection from OD over prolonged time scales [29,64,65]. The fact that less robust changes are observed on OD mortality caused by stimulants without opioids may be due to aforementioned lack of specific antidotes for stimulant OD, or due to potential differences in the populations who self-expose to opioids versus stimulants.

We and others have shown robust sex differences in synthetic opioid or stimulant OD, where males have greater mortality across jurisdictions and environments[6,9]. In this study, males had approximately 2-fold greater polydrug mortality than females overall, even across dynamic yearly trends in 2018-2024. Intriguingly, whereas rates of polydrug OD mortality differed in males versus females, an adjusted analysis did not detect sex as a significant predictor of *change* across years (i.e., APC in 2024 versus 2023), indicating that recent decreases in mortality did not differ in males versus females in this state-level analysis. However, the marginal adjusted p-value for sex (p=0.075) does suggest that future studies on this issue are warranted. In this adjusted analysis, we also found that the rate of prescribed naloxone dispensing[49] was not a significant predictor of these changes in the polydrug OD (of note these data did not include over-the-counter naloxone sales, to be examined in the future)[27,66]. Population-based data do indicate that large-scale distribution of intranasal naloxone can result in decreased OD [67,68]. Furthermore, it cannot be excluded that some layperson-administered naloxone reversals may go unreported [69], resulting in a potential underestimate of the impact of this intervention on OD mortality. Importantly, the adjusted analysis in this study did detect that a lesser decrease in these polydrug OD occurred in states that had a greater percentage of the population without health insurance, adjusting for household income. This result suggests that availability of continuing care is associated with better public health outcomes, even for acute emergencies such as OD.

Synthetic opioid or stimulant OD mortality has also been shown to differ by age, with peak mortality typically observed in mid-adulthood[6,41]. Overall, this study provides the first age stratification for national polydrug OD across 2018-2024, and shows large differences in mortality across the lifespan, with highest risk at ages 35-44, similar to a prior study for opioids and stimulants studied separately [6]. Nevertheless recent decreases in polydrug OD rates were observed even for this high-risk age group, based on joinpoint regressions. The reasons for this age-related profile in OD mortality are unclear, but may include the typical lifespan trajectory of illicit opioid or stimulant exposure, usually commencing in late adolescence and early adulthood, followed by years of greater disease severity and peak use [70], consistent with the mean age of persons presenting to emergency departments for such OD care [5,10]. Data in CDC WONDER do not provide the longer-term history of drug exposure prior to the lethal OD, therefore future studies have to explore underlying mechanisms [71] for the age distribution in this OD mortality. Intriguingly, impulsivity and risk-taking in the general population are reported to peak earlier in the lifespan (e.g., prior to age 25) [72,73], therefore these neurocognitive features alone are not necessarily the main underlying factor for the observed peak ages for polydrug OD mortality.

Race is an important social determinant of health, including for risk of OD mortality[6,48]. In a follow-up, we found that while polydrug OD rates differed across persons in the African-American/Black racial group versus those in the white group, there were similar annual percent changes (i.e., decreases) in 2024 versus 2023. This suggests that effective prevention, public health strategies and cascades of care can be effective across a broad range of communities and sociodemographic features [74,75].

Illicit drug markets and OD epidemiology can differ across countries [76]. For example, recent data from Canada (for 2018-2022) also find that fentanyl combinations with either cocaine or methamphetamine are the most common polydrug combinations observed in OD mortality [77]. However, it is unclear if a similar decrease in either opioid or stimulant OD (or their polydrug combination) has occurred in 2024 versus 2023 in Canada [77]. As synthetic opioids are potentially entering the illicit supply in several countries (e.g., in Europe and Australia), continued vigilance and localized public health interventions will be critical in the coming years [76,78,79].

### Limitations

This national study from the UnIted States has considerable strengths, but some limitations need to be considered.

**A)** The ICD-10 category for synthetic opioids is thought to reflect primarily fentanyl and its analogs[4], but can also contain MOR-agonists from other chemical scaffolds, including nitazenes[44]. Also, we did not study other types of MOR-agonists available for analysis in CDC WONDER (e.g., heroin, methadone or prescription opioids), as these are causing OD at a substantially lower rate than synthetic opioids, in recent years[6,80]. **B)** The “psychostimulants with abuse potential” ICD-10 category may primarily reflect methamphetamine, but presence of other amphetamine analogs or stimulants cannot be excluded. Also, for simplicity, we examined compounds such as methamphetamine and cocaine together, but their pathophysiological effects are not necessarily identical[81,82]. **C)** The data for 2024 are considered provisional at the time of this submission. However, there is relatively high completeness in CDC WONDER data after 6 months of occurrence [83], and the data for this study were obtained after such an interval (i.e., in June 2025). There may also be potential under-reporting or mis-reporting of OD mortality in death certificates[84,85]. **D)** For simplicity and based on the categories available in CDC WONDER, we did not examine further polydrug patterns (e.g., including benzodiazepines, alcohol or xylazine)[86–88]. Future studies should determine whether some of these other substances are particularly associated with the synthetic opioid and/or stimulant OD yearly trajectories in national data.

### Implications and conclusions

This study provides sex-specific national trajectories of mortality for polydrug OD caused by synthetic opioids and stimulants in 2018-2024, relevant to the national OD crisis. Overall, relatively large decreases in this polydrug OD mortality were detected in 2024 versus 2023, similarly for males and females. However, joinpoint regressions indicate that trajectories of polydrug OD mortality across the broader 2018-2024 time period were not identical across sex, for reasons that should be examined.

Furthermore, we also found that OD mortality caused by opioids *without* stimulants underwent a similar recent decrease in 2024, consistent with recent reports[41]. However, considerably smaller changes occurred in OD caused by stimulants *without* synthetic opioids. In addition, our age stratification analyses also show that while decreases in polydrug OD mortality were observed across a broad age range (overall range 15-74), persons in mid-adulthood remain at highest risk[70,89,90]. Overall, despite these recent decreases, polydrug OD rates in 2024 are still greater than those for the two other groups studied here, indicating that development of targeted prevention and intervention strategies is a continuing need[13,20,91]

## Supporting information

Supplementary Results

## Abbreviations

95%CI: 95% confidence intervals
AA: African American or Black
APC: annual percent change in rate of mortality; ((Rate_Year N_ - Rate_Year N-1_)/Rate_Year N-1_) X100%)
CDC: United States Centers for Disease Control and Prevention
F: Female
M: Male
MOR: mu-opioid receptor
MOUD: medications for opioid use disorder
NS: not significant
OD: overdose
SE: standard error

## Author Contributions

### Concept and Design: ERB

**Data acquisition:** publicly available de-identified data from CDC WONDER

**Analysis:** ERB

**Interpretation and writing:** All authors

### Funding

The authors gratefully acknowledge funding from NIDA-NIH DA5U01DA053625 (ERB), 2R01DA048009 (AM), 1R01DA060914 (RZG), 5R01DA049547 (NAK, RZG). The funder had no role in the design, drafting or editing of the project.

### Data Availability Statement

Overdose data analyzed herein are publicly available from the United States Centers for Disease Control and Prevention CDC WONDER platform (https://wonder.cdc.gov/mcd-icd10-provisional.html).

Naloxone dispensing rates from the CDC are publicly available from https://www.cdc.gov/overdose-prevention/data-research/facts-stats/naloxone-dispensing-rate-maps.html. Sociodemographic data are publicly available from the American Community Survey for 2023, from the United States Census Bureau (https://data.census.gov/)

## References

1. Han B, Einstein EB, Jones CM, Cotto J, Compton WM, Volkow ND. Racial and Ethnic Disparities in Drug Overdose Deaths in the US During the COVID-19 Pandemic. JAMA Netw Open. 2022;5: e2232314. doi:10.1001/jamanetworkopen.2022.32314

2. Brown KG, Chen CY, Dong D, Lake KJ, Butelman ER. Has the United States Reached a Plateau in Overdoses Caused by Synthetic Opioids After the Onset of the COVID-19 Pandemic? Examination of Centers for Disease Control and Prevention Data to November 2021. Front Psychiatry. 2022;https://www.frontiersin.org/articles/10.3389/fpsyt.2022.947603/full. Available: https://www.frontiersin.org/articles/10.3389/fpsyt.2022.947603/full

3. Ciccarone D. Bending the overdose curve — still not enough. N Engl J Med. 2024;391: 1052–1053. doi:10.1056/nejme2406359

4. Amaducci A, Aldy K, Campleman SL, Li S, Meyn A, Abston S, et al. Naloxone Use in Novel Potent Opioid and Fentanyl Overdoses in Emergency Department Patients. JAMA Netw Open. 2023;6: e2331264. doi:10.1001/jamanetworkopen.2023.31264

5. Shastry S, Lin J, Aldy K, Brent J, Wax P, Krotulski A, et al. Heroin or fentanyl: Prevalence of confirmed fentanyl in ED patients with suspected heroin overdose. J Am Coll Emerg Physicians Open. 2024;5: e13235. doi:10.1002/emp2.13235

6. Butelman ER, Huang Y, Epstein DH, Shaham Y, Goldstein RZ, Volkow ND, et al. Overdose mortality rates for opioids and stimulant drugs are substantially higher in men than in women: state-level analysis. Neuropsychopharmacology. 2023. doi:10.1038/s41386-023-01601-8

7. Ciccarone D. The rise of illicit fentanyls, stimulants and the fourth wave of the opioid overdose crisis. Curr Opin Psychiatry. 2021;34: 344–350. doi:10.1097/yco.0000000000000717

8. Tanz LJ, Dinwiddie AT, Mattson CL, O’Donnell J, Davis NL. Drug Overdose Deaths Among Persons Aged 10-19 Years - United States, July 2019-December 2021. MMWR Morb Mortal Wkly Rep. 2022;71: 1576–1582. doi:10.15585/mmwr.mm7150a2

9. Wilson N, Kariisa M, Seth P, Smith H 4th, Davis NL. Drug and Opioid-Involved Overdose Deaths - United States, 2017-2018. MMWR Morb Mortal Wkly Rep. 2020;69: 290–297. doi:10.15585/mmwr.mm6911a4

10. Shastry S, Shulman J, Aldy K, Brent J, Wax P, Manini AF, et al. Psychostimulant drug co-ingestion in non-fatal opioid overdose. Drug Alcohol Depend Rep. 2024;10: 100223. doi:10.1016/j.dadr.2024.100223

11. CDC. The Fentalog Study: A Subset of Nonfatal Suspected Opioid-Involved Overdoses with Toxicology Testing. In: Overdose Prevention [Internet]. 11 Oct 2024 [cited 20 Jun 2025]. Available: https://www.cdc.gov/overdose-prevention/data-research/facts-stats/fentalog-study-dashboard.html

12. Liu S, Scholl L, Hoots B, Seth P. Nonfatal drug and polydrug overdoses treated in emergency departments - 29 states, 2018-2019. MMWR Morb Mortal Wkly Rep. 2020;69: 1149–1155. doi:10.15585/mmwr.mm6934a1

13. Cotten JF. Goofball polypharmacy. J Pharmacol Exp Ther. 2024;388: 241–243. doi:10.1124/jpet.123.001930

14. Ondocsin J, Holm N, Mars SG, Ciccarone D. The motives and methods of methamphetamine and “heroin” co-use in West Virginia. Harm Reduct J. 2023;20: 88. doi:10.1186/s12954-023-00816-8

15. Volkow ND, Califf RM, Sokolowska M, Tabak LA, Compton WM. Testing for Fentanyl - Urgent Need for Practice-Relevant and Public Health Research. N Engl J Med. 2023. doi:10.1056/NEJMp2302857

16. Palamar JJ, Fitzgerald N, Carr TH, Cottler LB, Ciccarone D. National and regional trends in fentanyl seizures in the United States, 2017–2023. International Journal of Drug Policy. 2024; 104417. doi:10.1016/j.drugpo.2024.104417

17. Karandinos G, Unick J, Ciccarone D. Mortality risk among individuals who smoke opioids compared with those who inject: A propensity score-matched cohort analysis of United States national treatment data. Addiction. 2025. doi:10.1111/add.16740

18. Glidden E, Gladden RM, Dion C, Spyres MB, Seth P, Aldy K, et al. Suspected counterfeit M-30 oxycodone pill exposures and acute withdrawals reported from a single hospital - Toxicology Investigators Consortium Core Registry, U.s. census Bureau Western Region, 2017-2022. MMWR Morb Mortal Wkly Rep. 2024;73: 642–647. doi:10.15585/mmwr.mm7329a2

19. Allen C, Arredondo C, Dunham R, Fishman M, Lev L, Mace S, et al. Guidance for Handling the Increasing Prevalence of Drugs Adulterated or Laced With Fentanyl. PS. 2023;74: 1059–1062. doi:10.1176/appi.ps.202100660

20. Friedman J, Ciccarone D. The public health risks of counterfeit pills. Lancet Public Health. 2025;10: e58– e62. doi:10.1016/S2468-2667(24)00273-1

21. Skolnick P. Treatment of overdose in the synthetic opioid era. Pharmacol Ther. 2021; 108019. doi:10.1016/j.pharmthera.2021.108019

22. Baldo BA. Opioid-induced respiratory depression: clinical aspects and pathophysiology of the respiratory network effects. Am J Physiol Lung Cell Mol Physiol. 2025;328: L267–L289. doi:10.1152/ajplung.00314.2024

23. Ritz MC, Lamb RJ, Goldberg SR, Kuhar MJ. Cocaine receptors on dopamine transporters are related to self-administration of cocaine. Science. 1987;237: 1219–1223. doi:10.1126/science.2820058

24. Sulzer D, Chen TK, Lau YY, Kristensen H, Rayport S, Ewing A. Amphetamine redistributes dopamine from synaptic vesicles to the cytosol and promotes reverse transport. J Neurosci. 1995;15: 4102–4108. doi:10.1523/jneurosci.15-05-04102.1995

25. Richards JR, Laurin EG. Methamphetamine toxicity. StatPearls. Treasure Island (FL): StatPearls Publishing; 2025. Available: https://www.ncbi.nlm.nih.gov/books/NBK430895/

26. Heard K, Palmer R, Zahniser NR. Mechanisms of acute cocaine toxicity. Open Pharmacol J. 2008;2: 70–78. doi:10.2174/1874143600802010070

27. FDA approves first over-the-counter naloxone nasal spray. In: U.S. Food and Drug Administration [Internet]. FDA; 9 Aug 2024 [cited 5 Jun 2025]. Available: https://www.fda.gov/news-events/press-announcements/fda-approves-first-over-counter-naloxone-nasal-spray

28. Bennett AS, Freeman R, Des Jarlais DC, Aronson ID. Reasons people who use opioids do not accept or carry no-cost naloxone: Qualitative interview study. JMIR Form Res. 2020;4: e22411. doi:10.2196/22411

29. Wakeman SE, Larochelle MR, Ameli O, Chaisson CE, McPheeters JT, Crown WH, et al. Comparative Effectiveness of Different Treatment Pathways for Opioid Use Disorder. JAMA Netw Open. 2020;3: e1920622. doi:10.1001/jamanetworkopen.2019.20622

30. Shastry S, Carpenter J, Krotulski A, Brent J, Wax P, Aldy K, et al. Disparities in treatment and referral after an opioid overdose among emergency department patients. JAMA Netw Open. 2025;8: e2518569. doi:10.1001/jamanetworkopen.2025.18569

31. Hooker SA, Starkey C, Bart G, Rossom RC, Kane S, Olson AW. Predicting buprenorphine adherence among patients with opioid use disorder in primary care settings. BMC Prim Care. 2024;25: 361. doi:10.1186/s12875-024-02609-9

32. Wakeman SE, McGovern S, Kehoe L, Kane MT, Powell EA, Casey SK, et al. Predictors of engagement and retention in care at a low-threshold substance use disorder bridge clinic. J Subst Abuse Treat. 2022;141: 108848. doi:10.1016/j.jsat.2022.108848

33. D’Onofrio G, O’Connor PG, Pantalon MV, Chawarski MC, Busch SH, Owens PH, et al. Emergency Department-Initiated Buprenorphine/Naloxone Treatment for Opioid Dependence: A Randomized Clinical Trial. JAMA. 2015;313: 1636–1644. doi:10.1001/jama.2015.3474

34. National Institute on Drug Abuse. Reduced drug use is a meaningful treatment outcome for people with stimulant use disorders. In: National Institute on Drug Abuse [Internet]. 10 Jan 2024 [cited 29 Sep 2025]. Available: https://nida.nih.gov/news-events/news-releases/2024/01/reduced-drug-use-is-a-meaningful-treatment-outcome-for-people-with-stimulant-use-disorders

35. Hiranita T, Ho NP, France CP. Ventilatory effects of fentanyl, heroin, and d -methamphetamine, alone and in mixtures, in male rats breathing normal air. J Pharmacol Exp Ther. 2023. doi:10.1124/jpet.123.001653

36. Burke O, Ely SF, Gill JR. Cardiovascular disease in acute cocaine compared to acute fentanyl intoxication deaths. Am J Forensic Med Pathol. 2024. doi:10.1097/paf.0000000000000994

37. Zuin M, Mohanty S, Aggarwal R, Bertini M, Bikdeli B, Hamade N, et al. Trends in sudden cardiac death among adults aged 25 to 44 years in the United States: An analysis of 2 large US databases. J Am Heart Assoc. 2025;14: e035722. doi:10.1161/JAHA.124.035722

38. Mann J, Samieegohar M, Chaturbedi A, Zirkle J, Han X, Ahmadi SF, et al. Development of a Translational Model to Assess the Impact of Opioid Overdose and Naloxone Dosing on Respiratory Depression and Cardiac Arrest. Clin Pharmacol Ther. 2022;112: 1020–1032. doi:10.1002/cpt.2696

39. Reddy PKV, Ng TMH, Oh EE, Moady G, Elkayam U. Clinical characteristics and management of methamphetamine-associated cardiomyopathy: State-of-the-art review. J Am Heart Assoc. 2020;9: e016704. doi:10.1161/JAHA.120.016704

40. Kevil CG, Goeders NE, Woolard MD, Bhuiyan MS, Dominic P, Kolluru GK, et al. Methamphetamine Use and Cardiovascular Disease. Arterioscler Thromb Vasc Biol. 2019;39: 1739–1746. doi:10.1161/ATVBAHA.119.312461

41. Post LA, Ciccarone D, Unick GJ, D’Onofrio G, Kwon S, Lundberg AL, et al. Decline in US drug overdose deaths by region, substance, and demographics. JAMA Netw Open. 2025;8: e2514997. doi:10.1001/jamanetworkopen.2025.14997

42. Kiang MV, Humphreys K. Recent drug overdose mortality decline compared with pre-COVID-19 trend. JAMA Netw Open. 2025;8: e2458090. doi:10.1001/jamanetworkopen.2024.58090

43. Spencer MR, Garnett M, Miniño A. Drug overdose deaths in the United States, 2002-2022. National Center for Health Statistics (U.S.); 2023 Dec. doi:10.15620/cdc:135849

44. Berger JC, Severe AD, Jalloh MS, Manini AF. Naloxone dosing and hospitalization for nitazene overdose: A scoping review. J Med Toxicol. 2025. doi:10.1007/s13181-025-01059-8

45. Joinpoint Regression Program. [cited 10 Jun 2025]. Available: https://surveillance.cancer.gov/joinpoint

46. Ingram DD, Malec DJ, Makuc DM, Kruszon-Moran D, Gindi RM, Albert M, et al. National Center for Health Statistics guidelines for analysis of trends. Vital Health Stat 2. 2018; 1–71. Available: https://www.ncbi.nlm.nih.gov/pubmed/29775435

47. Kim H-J, Chen H-S, Byrne J, Wheeler B, Feuer EJ. Twenty years since Joinpoint 1.0: Two major enhancements, their justification, and impact. Stat Med. 2022;41: 3102–3130. doi:10.1002/sim.9407

48. Social Determinants of Health. [cited 12 Feb 2025]. Available: https://odphp.health.gov/healthypeople/priority-areas/social-determinants-health

49. CDC. Naloxone dispensing rate maps. In: Overdose Prevention [Internet]. 7 Nov 2024 [cited 5 Jun 2025]. Available: https://www.cdc.gov/overdose-prevention/data-research/facts-stats/naloxone-dispensing-rate-maps.html

50. U.S. Census Bureau. Explore Census Data. [cited 16 Jun 2025]. Available: https://data.census.gov/advanced

51. Hedegaard H, Miniño AM, Warner M. Co-involvement of opioids in drug overdose deaths involving cocaine and psychostimulants; 10.15620/cdc:103966. In: NCHS Data Brief, no 406. Hyattsville, MD: National Center for Health Statistics. 2021 [Internet]. https://doi.org/10.15620/cdc:103966; 2021. doi:10.15620/cdc:103966

52. Oles WC, Liu M, Wakeman SE, Larochelle MR. Geographic trends in opioid and polysubstance overdose deaths in the US, 2014–2023. J Gen Intern Med. 2025. doi:10.1007/s11606-025-09589-1

53. Volkow N, Dye LR. Groundbreaking research from NIDA addressing the challenges of the opioid epidemic. J Med Toxicol. 2025;21: 69–77. doi:10.1007/s13181-024-01041-w

54. Levengood TW, Shaw LC, Zang X, Schackman BR, Walley AY, Urquhart C, et al. A geospatial analysis of naloxone distribution patterns in the Massachusetts overdose education and naloxone distribution program (MA OEND). Drug Alcohol Depend. 2025;274: 112783. doi:10.1016/j.drugalcdep.2025.112783

55. Drug Enforcement Administration: 2025 National Drug Threat Assessment. [cited 8 Oct 2025]. Available: https://www.dea.gov/sites/default/files/2025-07/2025NationalDrugThreatAssessment.pdf

56. Naloxone Distribution Project. In: California Department of Health Care Services [Internet]. [cited 1 Oct 2025]. Available: https://www.dhcs.ca.gov/individuals/Pages/Naloxone_Distribution_Project.aspx

57. OASAS announces the availability of lifesaving resources to address the state’s opioid crisis. In: Office of Addiction Services and Supports [Internet]. [cited 1 Oct 2025]. Available: https://oasas.ny.gov/news/oasas-announces-availability-lifesaving-resources-address-states-opioid-crisis

58. Gelberg KH, El-Bassel N, Babineau DC, Vickers-Smith RA, Fanucchi LC, Childerhose JE, et al. Association of fentanyl test strip results and change in drug use behaviors: A multi-state, community-based observational study. Int J Drug Policy. 2025;143: 104867. doi:10.1016/j.drugpo.2025.104867

59. Park JN, Tardif J, Thompson E, Rosen JG, Lira JAS, Green TC. A survey of North American drug checking services operating in 2022. Int J Drug Policy. 2023;121: 104206. doi:10.1016/j.drugpo.2023.104206

60. Maghsoudi N, Tanguay J, Scarfone K, Rammohan I, Ziegler C, Werb D, et al. Drug checking services for people who use drugs: a systematic review. Addiction. 2022;117: 532–544. doi:10.1111/add.15734

61. Samuels EA, Rosen AD, Abusaa S, Dekker AM, Schriger DL, Shoptaw SJ, et al. Increasing emergency department patient navigation and buprenorphine use: A model for low-barrier treatment. Health Aff (Millwood). 2025;44: 1138–1147. doi:10.1377/hlthaff.2025.00333

62. Dekker AM, Schriger DL, Herring AA, Samuels EA. Emergency clinician buprenorphine initiation, subsequent prescriptions, and continuous prescriptions. JAMA. 2025. doi:10.1001/jama.2024.27976

63. Hughes T, Nasser N, Mitra A. Overview of best practices for buprenorphine initiation in the emergency department. Int J Emerg Med. 2024;17: 23. doi:10.1186/s12245-024-00593-6

64. Jones CM, Shoff C, Blanco C, Losby JL, Ling SM, Compton WM. Overdose, Behavioral Health Services, and Medications for Opioid Use Disorder After a Nonfatal Overdose. JAMA Intern Med. 2024. doi:10.1001/jamainternmed.2024.1733

65. Larochelle MR, Bernson D, Land T, Stopka TJ, Wang N, Xuan Z, et al. Medication for Opioid Use Disorder After Nonfatal Opioid Overdose and Association With Mortality: A Cohort Study. Ann Intern Med. 2018;169: 137–145. doi:10.7326/m17-3107

66. Marley GT, Annis IE, Egan KL, Delamater P, Carpenter DM. Naloxone Availability and Cost After Transition to an Over-the-Counter Product. JAMA Health Forum. 2024;5: e241920. doi:10.1001/jamahealthforum.2024.1920

67. Håkansson A, Alanko Blomé M, Isendahl P, Landgren M, Malmqvist U, Troberg K. Distribution of intranasal naloxone to potential opioid overdose bystanders in Sweden: effects on overdose mortality in a full region-wide study. BMJ Open. 2024;14: e074152. doi:10.1136/bmjopen-2023-074152

68. Walley AY, Xuan Z, Hackman HH, Quinn E, Doe-Simkins M, Sorensen-Alawad A, et al. Opioid overdose rates and implementation of overdose education and nasal naloxone distribution in Massachusetts: interrupted time series analysis. BMJ. 2013;346: f174. doi:10.1136/bmj.f174

69. Moustaqim-Barrette A, Papamihali K, Williams S, Ferguson M, Moe J, Purssell R, et al. Adverse events related to bystander naloxone administration in cases of suspected opioid overdose in British Columbia: An observational study. PLoS One. 2021;16: e0259126. doi:10.1371/journal.pone.0259126

70. Butelman ER, Chen CY, Brown KG, Kreek MJ. Escalation of drug use in persons dually diagnosed with opioid and cocaine dependence: Gender comparison and dimensional predictors. Drug Alcohol Depend. 2019;205: 107657. doi:10.1016/j.drugalcdep.2019.107657

71. Boitor M, Ballard A, Emed J, Le May S, Gélinas C. Risk factors for severe opioid-induced respiratory depression in hospitalized adults: A case-control study. Can J Pain. 2020;4: 103–110. doi:10.1080/24740527.2020.1714431

72. Duell N, Steinberg L, Icenogle G, Chein J, Chaudhary N, Di Giunta L, et al. Age patterns in risk taking across the world. J Youth Adolesc. 2018;47: 1052–1072. doi:10.1007/s10964-017-0752-y

73. Romer D. Adolescent risk taking, impulsivity, and brain development: implications for prevention. Dev Psychobiol. 2010;52: 263–276. doi:10.1002/dev.20442

74. McLellan AT, Volkow ND. Goals for opioid use disorder medications - protection, remission, and recovery. N Engl J Med. 2025. doi:10.1056/NEJMp2505377

75. Kariisa M, Davis NL, Kumar S, Seth P, Mattson CL, Chowdhury F, et al. Vital signs: Drug overdose deaths, by selected sociodemographic and social determinants of health characteristics - 25 states and the District of Columbia, 2019-2020. MMWR Morb Mortal Wkly Rep. 2022;71: 940–947. doi:10.15585/mmwr.mm7129e2

76. What are fentanyl and nitazenes? Explaining the rise and risks of potent synthetic opioids. [cited 1 Oct 2025]. Available: https://adf.org.au/insights/fentanyl-and-nitazenes/

77. Health Canada. Multi-drug combinations in national apparent opioid and stimulant toxicity deaths. 10 Jul 2024 [cited 1 Oct 2025]. Available: https://www.canada.ca/en/health-canada/services/opioids/data-surveillance-research/multi-drug-combinations-national-opioid-stimulant-toxicity-deaths.html

78. Roberts A-O, Richards GC. Is England facing an opioid epidemic? Br J Pain. 2023;17: 320–324. doi:10.1177/20494637231160684

79. European Drug Report 2025: Trends and Developments. [cited 1 Oct 2025]. Available: https://www.euda.europa.eu/publications/european-drug-report/2025_en

80. Jones CM, Compton WM, Han B, Baldwin G, Volkow ND. Methadone-Involved Overdose Deaths in the US Before and After Federal Policy Changes Expanding Take-Home Methadone Doses From Opioid Treatment Programs. JAMA Psychiatry. 2022 [cited 13 Jul 2022]. doi:10.1001/jamapsychiatry.2022.1776

81. Zimmerman JL. Cocaine intoxication. Crit Care Clin. 2012;28: 517–526. doi:10.1016/j.ccc.2012.07.003

82. Curran L, Nah G, Marcus GM, Tseng Z, Crawford MH, Parikh NI. Clinical Correlates and Outcomes of Methamphetamine-Associated Cardiovascular Diseases in Hospitalized Patients in California. J Am Heart Assoc. 2022;11: e023663. doi:10.1161/JAHA.121.023663

83. Spencer MR, Ahmad S. Timeliness of Death Certificate Data for Mortality Surveillance and Provisional Estimates. In: National Vital Statistics System [Internet]. 2016. Available: https://www.cdc.gov/nchs/data/vsrr/report001.pdf

84. Johnson SC, Cunningham M, Dippenaar IN, Sharara F, Wool EE, Agesa KM, et al. Public health utility of cause of death data: applying empirical algorithms to improve data quality. BMC Med Inform Decis Mak. 2021;21: 175. doi:10.1186/s12911-021-01501-1

85. Boslett AJ, Denham A, Hill EL. Using contributing causes of death improves prediction of opioid involvement in unclassified drug overdoses in US death records: Methods for predicting fatal opioid overdoses. Addiction. 2020;115: 1308–1317. doi:10.1111/add.14943

86. Love JS, Levine M, Aldy K, Brent J, Krotulski AJ, Logan BK, et al. Opioid overdoses involving xylazine in emergency department patients: a multicenter study. Clin Toxicol. 2023;61: 173–180. doi:10.1080/15563650.2022.2159427

87. Friedman JR. Assessing an ICD-10 code approach for estimating xylazine-involved overdose deaths in the United States. Drug Alcohol Depend. 2025;274: 112781. doi:10.1016/j.drugalcdep.2025.112781

88. Hughes A, Spungen H, Culbreth R, Aldy K, Krotulski A, Hendrickson RG, et al. Benzodiazepine co-exposure among patients presenting to the emergency department with a confirmed opioid overdose. Acad Emerg Med. 2025. doi:10.1111/acem.70104

89. Woodcock EA, Lundahl LH, Stoltman JJ, Greenwald MK. Progression to regular heroin use: examination of patterns, predictors, and consequences. Addict Behav. 2015;45: 287–293. doi:10.1016/j.addbeh.2015.02.014

90. Rodriguez-Cintas L, Daigre C, Grau-Lopez L, Barral C, Perez-Pazos J, Voltes N, et al. Impulsivity and addiction severity in cocaine and opioid dependent patients. Addict Behav. 2016;58: 104–109. doi:10.1016/j.addbeh.2016.02.029

91. Lim TY, Dong H, Stringfellow E, Hasgul Z, Park J, Glos L, et al. Temporal and spatial trends of fentanyl co-occurrence in the illicit drug supply in the United States: a serial cross-sectional analysis. Lancet Reg Health Am. 2024;39: 100898. doi:10.1016/j.lana.2024.100898

