## Supplementary Results for "Polydrug overdose mortality caused by synthetic opioids and stimulants: Current sex- and age- specific trajectories in United States national data for 2018-2024"

**Supplement**

**Table S1A:** National data for annual overdose (OD) mortality caused by polydrug synthetic opioids *and* stimulants /100,000 population

| **Sex** | **Year** | **Deaths** | **Population** | **Crude Rate** | **Age-Adjusted Rate** | **Age-Adjusted Rate SE** |
| --- | --- | --- | --- | --- | --- | --- |
| Female | 2018 | 3404 | 123382199 | 2.76 | 3.01 | 0.05 |
| Female | 2019 | 4116 | 123761464 | 3.33 | 3.63 | 0.06 |
| Female | 2020 | 6625 | 124265374 | 5.33 | 5.85 | 0.07 |
| Female | 2021 | 9782 | 125093692 | 7.82 | 8.49 | 0.09 |
| Female | 2022 | 10675 | 124987770 | 8.54 | 9.28 | 0.09 |
| Female | 2023 | 11164 | 125982434 | 8.86 | 9.55 | 0.09 |
| Female | 2024 (provisional) | 7269 | 125982434 | 5.77 | 6.21 | 0.07 |
| Male | 2018 | 8097 | 120960914 | 6.69 | 7.12 | 0.08 |
| Male | 2019 | 10493 | 121332383 | 8.65 | 9.18 | 0.09 |
| Male | 2020 | 17125 | 121815356 | 14.06 | 14.97 | 0.12 |
| Male | 2021 | 24588 | 124051552 | 19.82 | 21 | 0.14 |
| Male | 2022 | 28343 | 124855987 | 22.7 | 23.91 | 0.14 |
| Male | 2023 | 30279 | 124870903 | 24.25 | 25.36 | 0.15 |
| Male | 2024 (provisional) | 18552 | 124870903 | 14.86 | 15.57 | 0.12 |

**Table S1B:** National data for annual OD mortality caused by synthetic opioids *without* stimulants /100,000 population

| **Sex Code** | **Year** | **Deaths** | **Population** | **Crude Rate** | **Age-Adjusted Rate** | **Age-Adjusted Rate SE** |
| --- | --- | --- | --- | --- | --- | --- |
| Female | 2018 | 5354 | 123382199 | 4.34 | 4.59 | 0.09 |
| Female | 2019 | 5896 | 123761464 | 4.76 | 4.99 | 0.10 |
| Female | 2020 | 8525 | 124265374 | 6.86 | 7.31 | 0.12 |
| Female | 2021 | 9623 | 125093692 | 7.69 | 8.16 | 0.14 |
| Female | 2022 | 9032 | 124987770 | 7.23 | 7.61 | 0.14 |
| Female | 2023 | 8047 | 125982434 | 6.39 | 6.72 | 0.14 |
| Female | 2024 (provisional) | 4864 | 125982434 | 3.86 | 4.05 | 0.11 |
| Male | 2018 | 14364 | 120960914 | 11.88 | 12.48 | 0.14 |
| Male | 2019 | 15709 | 121332383 | 12.95 | 13.60 | 0.16 |
| Male | 2020 | 24012 | 121815356 | 19.71 | 20.70 | 0.20 |
| Male | 2021 | 26239 | 124051552 | 21.15 | 22.18 | 0.22 |
| Male | 2022 | 25289 | 124855987 | 20.26 | 21.21 | 0.23 |
| Male | 2023 | 22688 | 124870903 | 18.17 | 19.00 | 0.23 |
| Male | 2024 (provisional) | 13095 | 124870903 | 10.48 | 10.95 | 0.18 |

**Table S1C:** National data for annual OD mortality caused by stimulants *without* synthetic opioids /100,000 population

| **Sex Code** | **Year Code** | **Deaths** | **Population** | **Crude Rate** | **Age-Adjusted Rate** | **Age-Adjusted Rate SE** |
| --- | --- | --- | --- | --- | --- | --- |
| Female | 2018 | 4098 | 123382199 | 3.32 | 3.48 | 0.08 |
| Female | 2019 | 4390 | 123761464 | 3.54 | 3.69 | 0.09 |
| Female | 2020 | 4677 | 124265374 | 3.77 | 3.89 | 0.11 |
| Female | 2021 | 5251 | 125093692 | 4.2 | 4.29 | 0.13 |
| Female | 2022 | 5043 | 124987770 | 4.04 | 4.12 | 0.13 |
| Female | 2023 | 4857 | 125982434 | 3.86 | 3.84 | 0.13 |
| Female | 2024 (provisional) | 4619 | 125982434 | 3.67 | 3.64 | 0.11 |
| Male | 2018 | 10192 | 120960914 | 8.43 | 8.52 | 0.13 |
| Male | 2019 | 11128 | 121332383 | 9.17 | 9.28 | 0.15 |
| Male | 2020 | 12069 | 121815356 | 9.91 | 9.95 | 0.18 |
| Male | 2021 | 13661 | 124051552 | 11.01 | 10.91 | 0.20 |
| Male | 2022 | 13209 | 124855987 | 10.58 | 10.46 | 0.21 |
| Male | 2023 | 13079 | 124870903 | 10.47 | 10.14 | 0.22 |
| Male | 2024 (provisional) | 12361 | 124870903 | 9.9 | 9.58 | 0.18 |

**Table S2A:** Census division level data for annual OD mortality caused by polydrug synthetic opioids *and* stimulants/100,000 population (APC: annual percent change)

| **Sex** | **Year** | **Residence Census Division** | **Deaths** | **Population** | **Crude Rate** | **Age-Adjusted Rate** | **Age-adjusted APC 2024 vs 2023** |
| --- | --- | --- | --- | --- | --- | --- | --- |
| Female | 2023 | Division 1: New England | 652 | 5863205 | 11.12 | 12.25 | -25.55 |
| Female | 2023 | Division 2: Middle Atlantic | 1533 | 15891277 | 9.65 | 10.19 | -34.05 |
| Female | 2023 | Division 3: East North Central | 1797 | 17689240 | 10.16 | 11.2 | -41.61 |
| Female | 2023 | Division 4: West North Central | 464 | 7965262 | 5.83 | 6.49 | -33.59 |
| Female | 2023 | Division 5: South Atlantic | 2720 | 25978834 | 10.47 | 11.43 | -39.72 |
| Female | 2023 | Division 6: East South Central | 969 | 7477711 | 12.96 | 14.54 | -39.34 |
| Female | 2023 | Division 7: West South Central | 686 | 15660188 | 4.38 | 4.68 | -31.62 |
| Female | 2023 | Division 8: Mountain | 714 | 9517869 | 7.5 | 8.04 | -10.32 |
| Female | 2023 | Division 9: Pacific | 1629 | 19938848 | 8.17 | 8.41 | -25.33 |
| Female | 2024 | Division 1: New England | 483 | 5863205 | 8.24 | 9.12 |  |
| Female | 2024 | Division 2: Middle Atlantic | 1001 | 15891277 | 6.3 | 6.72 |  |
| Female | 2024 | Division 3: East North Central | 1054 | 17689240 | 5.96 | 6.54 |  |
| Female | 2024 | Division 4: West North Central | 312 | 7965262 | 3.92 | 4.31 |  |
| Female | 2024 | Division 5: South Atlantic | 1653 | 25978834 | 6.36 | 6.89 |  |
| Female | 2024 | Division 6: East South Central | 591 | 7477711 | 7.9 | 8.82 |  |
| Female | 2024 | Division 7: West South Central | 474 | 15660188 | 3.03 | 3.2 |  |
| Female | 2024 | Division 8: Mountain | 640 | 9517869 | 6.72 | 7.21 |  |
| Female | 2024 | Division 9: Pacific | 1208 | 19938848 | 6.06 | 6.28 |  |
| Male | 2023 | Division 1: New England | 1806 | 5727696 | 31.53 | 33.56 | -34.48 |
| Male | 2023 | Division 2: Middle Atlantic | 4338 | 15539560 | 27.92 | 28.7 | -40.94 |
| Male | 2023 | Division 3: East North Central | 4589 | 17613663 | 26.05 | 27.56 | -45.03 |
| Male | 2023 | Division 4: West North Central | 1031 | 8105544 | 12.72 | 13.67 | -36.28 |
| Male | 2023 | Division 5: South Atlantic | 6626 | 25075193 | 26.42 | 27.83 | -41.14 |
| Male | 2023 | Division 6: East South Central | 2376 | 7215788 | 32.93 | 36.08 | -43.93 |
| Male | 2023 | Division 7: West South Central | 1785 | 15613840 | 11.43 | 11.97 | -33.92 |
| Male | 2023 | Division 8: Mountain | 2091 | 9736548 | 21.48 | 22.61 | -22.51 |
| Male | 2023 | Division 9: Pacific | 5637 | 20243071 | 27.85 | 28.14 | -29.57 |
| Male | 2024 | Division 1: New England | 1182 | 5727696 | 20.64 | 21.99 |  |
| Male | 2024 | Division 2: Middle Atlantic | 2551 | 15539560 | 16.42 | 16.95 |  |
| Male | 2024 | Division 3: East North Central | 2523 | 17613663 | 14.32 | 15.15 |  |
| Male | 2024 | Division 4: West North Central | 659 | 8105544 | 8.13 | 8.71 |  |
| Male | 2024 | Division 5: South Atlantic | 3879 | 25075193 | 15.47 | 16.38 |  |
| Male | 2024 | Division 6: East South Central | 1339 | 7215788 | 18.56 | 20.23 |  |
| Male | 2024 | Division 7: West South Central | 1167 | 15613840 | 7.47 | 7.91 |  |
| Male | 2024 | Division 8: Mountain | 1626 | 9736548 | 16.7 | 17.52 |  |
| Male | 2024 | Division 9: Pacific | 3968 | 20243071 | 19.6 | 19.82 |  |

**Table S2B:** Census division level data for annual OD mortality caused by synthetic opioids *without* stimulants/100,000 population

| **Sex** | **Year** | **Residence Census Division** | **Deaths** | **Population** | **Crude Rate** | **Age-Adjusted Rate** | **Age-adjusted APC 2024 vs 2023** |
| --- | --- | --- | --- | --- | --- | --- | --- |
| Female | 2023 | Division 1: New England | 514 | 5863205 | 8.77 | 9.24 | -42.21 |
| Female | 2023 | Division 2: Middle Atlantic | 1150 | 15891277 | 7.23 | 7.46 | -43.3 |
| Female | 2023 | Division 3: East North Central | 1330 | 17689240 | 7.52 | 8 | -40.63 |
| Female | 2023 | Division 4: West North Central | 419 | 7965262 | 5.26 | 5.72 | -40.91 |
| Female | 2023 | Division 5: South Atlantic | 1917 | 25978834 | 7.38 | 7.85 | -37.2 |
| Female | 2023 | Division 6: East South Central | 688 | 7477711 | 9.2 | 10.1 | -37.92 |
| Female | 2023 | Division 7: West South Central | 619 | 15660188 | 3.95 | 4.09 | -34.96 |
| Female | 2023 | Division 8: Mountain | 529 | 9517869 | 5.56 | 5.7 | -32.11 |
| Female | 2023 | Division 9: Pacific | 881 | 19938848 | 4.42 | 4.52 | -36.95 |
| Female | 2024 | Division 1: New England | 301 | 5863205 | 5.13 | 5.34 |  |
| Female | 2024 | Division 2: Middle Atlantic | 659 | 15891277 | 4.15 | 4.23 |  |
| Female | 2024 | Division 3: East North Central | 795 | 17689240 | 4.49 | 4.75 |  |
| Female | 2024 | Division 4: West North Central | 251 | 7965262 | 3.15 | 3.38 |  |
| Female | 2024 | Division 5: South Atlantic | 1195 | 25978834 | 4.6 | 4.93 |  |
| Female | 2024 | Division 6: East South Central | 433 | 7477711 | 5.79 | 6.27 |  |
| Female | 2024 | Division 7: West South Central | 397 | 15660188 | 2.53 | 2.66 |  |
| Female | 2024 | Division 8: Mountain | 362 | 9517869 | 3.81 | 3.87 |  |
| Female | 2024 | Division 9: Pacific | 558 | 19938848 | 2.8 | 2.85 |  |
| Male | 2023 | Division 1: New England | 1533 | 5727696 | 26.77 | 28.13 | -46.57 |
| Male | 2023 | Division 2: Middle Atlantic | 3360 | 15539560 | 21.62 | 22 | -41.86 |
| Male | 2023 | Division 3: East North Central | 3483 | 17613663 | 19.78 | 20.79 | -43.87 |
| Male | 2023 | Division 4: West North Central | 1074 | 8105544 | 13.25 | 14.24 | -44.59 |
| Male | 2023 | Division 5: South Atlantic | 5119 | 25075193 | 20.42 | 21.56 | -42.86 |
| Male | 2023 | Division 6: East South Central | 1721 | 7215788 | 23.85 | 26.11 | -42.74 |
| Male | 2023 | Division 7: West South Central | 1532 | 15613840 | 9.81 | 10.18 | -36.25 |
| Male | 2023 | Division 8: Mountain | 1603 | 9736548 | 16.46 | 17.35 | -40.46 |
| Male | 2023 | Division 9: Pacific | 3263 | 20243071 | 16.12 | 16.54 | -35.13 |
| Male | 2024 | Division 1: New England | 834 | 5727696 | 14.56 | 15.03 | **N/A** |
| Male | 2024 | Division 2: Middle Atlantic | 1987 | 15539560 | 12.78 | 12.79 |  |
| Male | 2024 | Division 3: East North Central | 1958 | 17613663 | 11.12 | 11.67 |  |
| Male | 2024 | Division 4: West North Central | 596 | 8105544 | 7.35 | 7.89 |  |
| Male | 2024 | Division 5: South Atlantic | 2931 | 25075193 | 11.69 | 12.32 |  |
| Male | 2024 | Division 6: East South Central | 988 | 7215788 | 13.69 | 14.95 |  |
| Male | 2024 | Division 7: West South Central | 971 | 15613840 | 6.22 | 6.49 |  |
| Male | 2024 | Division 8: Mountain | 958 | 9736548 | 9.84 | 10.33 |  |
| Male | 2024 | Division 9: Pacific | 2117 | 20243071 | 10.46 | 10.73 |  |

**Table S2C:** Census division level data for annual OD mortality caused by stimulants *without* synthetic opioids /100,000 population

| **Sex** | **Year** | **Residence Census Division** | **Deaths** | **Population** | **Crude Rate** | **Age-Adjusted Rate** | **Age-adjusted APC 2024 vs 2023** |
| --- | --- | --- | --- | --- | --- | --- | --- |
| Female | 2023 | Division 1: New England | 132 | 5863205 | 2.25 | 2.29 | 0.44 |
| Female | 2023 | Division 2: Middle Atlantic | 423 | 15891277 | 2.66 | 2.51 | 0.8 |
| Female | 2023 | Division 3: East North Central | 589 | 17689240 | 3.33 | 3.37 | -7.12 |
| Female | 2023 | Division 4: West North Central | 261 | 7965262 | 3.27 | 3.33 | 9.31 |
| Female | 2023 | Division 5: South Atlantic | 863 | 25978834 | 3.32 | 3.33 | -5.71 |
| Female | 2023 | Division 6: East South Central | 380 | 7477711 | 5.08 | 5.29 | -9.45 |
| Female | 2023 | Division 7: West South Central | 718 | 15660188 | 4.59 | 4.68 | -1.71 |
| Female | 2023 | Division 8: Mountain | 519 | 9517869 | 5.45 | 5.53 | 0.72 |
| Female | 2023 | Division 9: Pacific | 972 | 19938848 | 4.87 | 4.7 | -4.68 |
| Female | 2024 | Division 1: New England | 129 | 5863205 | 2.2 | 2.3 | **N/A** |
| Female | 2024 | Division 2: Middle Atlantic | 425 | 15891277 | 2.67 | 2.53 |  |
| Female | 2024 | Division 3: East North Central | 544 | 17689240 | 3.07 | 3.13 |  |
| Female | 2024 | Division 4: West North Central | 275 | 7965262 | 3.45 | 3.64 |  |
| Female | 2024 | Division 5: South Atlantic | 801 | 25978834 | 3.09 | 3.14 |  |
| Female | 2024 | Division 6: East South Central | 340 | 7477711 | 4.55 | 4.79 |  |
| Female | 2024 | Division 7: West South Central | 719 | 15660188 | 4.59 | 4.6 |  |
| Female | 2024 | Division 8: Mountain | 528 | 9517869 | 5.55 | 5.57 |  |
| Female | 2024 | Division 9: Pacific | 940 | 19938848 | 4.71 | 4.48 |  |
| Male | 2023 | Division 1: New England | 306 | 5727696 | 5.34 | 5.22 | 1.53 |
| Male | 2023 | Division 2: Middle Atlantic | 1125 | 15539560 | 7.24 | 6.83 | -2.64 |
| Male | 2023 | Division 3: East North Central | 1486 | 17613663 | 8.44 | 8.31 | -4.57 |
| Male | 2023 | Division 4: West North Central | 690 | 8105544 | 8.51 | 8.48 | -6.84 |
| Male | 2023 | Division 5: South Atlantic | 2194 | 25075193 | 8.75 | 8.43 | -9.61 |
| Male | 2023 | Division 6: East South Central | 849 | 7215788 | 11.76 | 11.98 | -4.26 |
| Male | 2023 | Division 7: West South Central | 2214 | 15613840 | 14.18 | 14.17 | -7.76 |
| Male | 2023 | Division 8: Mountain | 1341 | 9736548 | 13.77 | 13.64 | 9.24 |
| Male | 2023 | Division 9: Pacific | 2874 | 20243071 | 14.19 | 13.38 | -1.49 |
| Male | 2024 | Division 1: New England | 312 | 5727696 | 5.44 | 5.3 | **N/A** |
| Male | 2024 | Division 2: Middle Atlantic | 1098 | 15539560 | 7.06 | 6.65 |  |
| Male | 2024 | Division 3: East North Central | 1422 | 17613663 | 8.08 | 7.93 |  |
| Male | 2024 | Division 4: West North Central | 641 | 8105544 | 7.91 | 7.9 |  |
| Male | 2024 | Division 5: South Atlantic | 1990 | 25075193 | 7.94 | 7.62 |  |
| Male | 2024 | Division 6: East South Central | 821 | 7215788 | 11.37 | 11.47 |  |
| Male | 2024 | Division 7: West South Central | 2048 | 15613840 | 13.12 | 13.07 |  |
| Male | 2024 | Division 8: Mountain | 1474 | 9736548 | 15.14 | 14.9 |  |
| Male | 2024 | Division 9: Pacific | 2834 | 20243071 | 14 | 13.18 |  |

**Table S3:** State-level age-adjusted rates for OD mortality caused by polydrug synthetic opioids *and* stimulants / 100,000 population for 2023-2024

| **State** | **Female** | | | **Male** | | |
| --- | --- | --- | --- | --- | --- | --- |
|  | **OD Rate 2023** | **OD Rate 2024** | **APC** | **OD Rate 2023** | **OD Rate 2024** | **APC** |
| Alabama | 10.08 | 6 | -40.48 | 23.7 | 16.61 | -29.92 |
| Alaska | 21.97 | 19.45 | -11.47 | 38.17 | 40.03 | 4.87 |
| Arizona | 8.94 | 9.28 | 3.8 | 29.67 | 23.95 | -19.28 |
| Arkansas | 3.6 | 2.58 | -28.33 | 8.49 | 4.55 | -46.41 |
| California | 6.68 | 4.24 | -36.53 | 24.25 | 15 | -38.14 |
| Colorado | 7.74 | 5.78 | -25.32 | 21.69 | 14.17 | -34.67 |
| Connecticut | 9.41 | 9.85 | 4.68 | 34.28 | 21.81 | -36.38 |
| Delaware | 26.14 | 14.38 | -44.99 | 47.1 | 25.59 | -45.67 |
| District of Columbia | 22.88 | 14.45 | -36.84 | 50.38 | 34.29 | -31.94 |
| Florida | 8.51 | 5.44 | -36.08 | 22.8 | 13.54 | -40.61 |
| Georgia | 7.85 | 4.69 | -40.25 | 16.78 | 11.01 | -34.39 |
| Idaho | 4.43 | 3.63 | -18.06 | 10.08 | 5.86 | -41.87 |
| Illinois | 7.46 | 4.5 | -39.68 | 22.78 | 12.92 | -43.28 |
| Indiana | 12.18 | 7.81 | -35.88 | 24.76 | 14.71 | -40.59 |
| Iowa | 3.81 | 2.44 | -35.96 | 6.01 | 3.51 | -41.6 |
| Kansas | 5.65 | 5.41 | -4.25 | 12.33 | 7.89 | -36.01 |
| Kentucky | 16.75 | 9.58 | -42.81 | 38.81 | 20.97 | -45.97 |
| Louisiana | 9.83 | 5.11 | -48.02 | 23.61 | 14.65 | -37.95 |
| Maine | 15.27 | 15.52 | 1.64 | 51.54 | 34.23 | -33.59 |
| Maryland | 11.63 | 6.36 | -45.31 | 31.23 | 17.96 | -42.49 |
| Massachusetts | 13.14 | 7.92 | -39.73 | 32.75 | 20.11 | -38.6 |
| Michigan | 9.94 | 5.62 | -43.46 | 24.05 | 11.52 | -52.1 |
| Minnesota | 7.73 | 4.69 | -39.33 | 17.79 | 10.39 | -41.6 |
| Mississippi | 6.3 | 4.37 | -30.63 | 18.26 | 9.74 | -46.66 |
| Missouri | 9.89 | 5.64 | -42.97 | 20.23 | 13.42 | -33.66 |
| Nevada | 9 | 9.34 | 3.78 | 28.01 | 25.7 | -8.25 |
| New Jersey | 8.47 | 5.65 | -33.29 | 25.28 | 17.15 | -32.16 |
| New Mexico | 16.64 | 11.57 | -30.47 | 36.85 | 23.41 | -36.47 |
| New York | 10.23 | 6.12 | -40.18 | 29.71 | 17.05 | -42.61 |
| North Carolina | 13.81 | 9.53 | -30.99 | 33.53 | 21.08 | -37.13 |
| Ohio | 15.76 | 8.9 | -43.53 | 37.31 | 20.01 | -46.37 |
| Oklahoma | 8.62 | 6.09 | -29.35 | 20.82 | 13.08 | -37.18 |
| Oregon | 12.05 | 10.12 | -16.02 | 41.2 | 29.65 | -28.03 |
| Pennsylvania | 11.52 | 8.06 | -30.03 | 29.83 | 15.69 | -47.4 |
| Rhode Island | 17.73 | 8.53 | -51.89 | 33.5 | 23.84 | -28.84 |
| South Carolina | 14.42 | 7.41 | -48.61 | 35.05 | 18.82 | -46.31 |
| Tennessee | 19.73 | 11.64 | -41 | 49.94 | 25.4 | -49.14 |
| Texas | 3.55 | 2.53 | -28.73 | 9.58 | 6.5 | -32.15 |
| Utah | 4.54 | 3.67 | -19.16 | 10.14 | 8.74 | -13.81 |
| Virginia | 10.72 | 5.88 | -45.15 | 26.86 | 13.21 | -50.82 |
| Washington | 14.95 | 12.84 | -14.11 | 42.68 | 34.84 | -18.37 |
| West Virginia | 40.01 | 17.87 | -55.34 | 82.74 | 42.48 | -48.66 |
| Wisconsin | 11.38 | 5.3 | -53.43 | 27.86 | 14.95 | -46.34 |

*Other states were excluded due to data suppression or unreliable estimates. APC: annual percent rate

**Table S4:** Socio-demographic and public health variables: State-level data for 2023

| State | Naloxone Dispensing Rate / 100 population^a^ | Median household income ($) | No health insurance coverage (%) | %African-American Population |
| --- | --- | --- | --- | --- |
| Alabama | 0.40 | 62212 | 8.5 | 25.4 |
| Alaska | 0.50 | 86631 | 10.4 | 2.9 |
| Arizona | 0.70 | 77315 | 9.9 | 4.8 |
| Arkansas | 1.90 | 58700 | 8.9 | 14.4 |
| California | 0.70 | 95521 | 6.4 | 5.4 |
| Colorado | 0.60 | 92911 | 6.7 | 3.9 |
| Connecticut | 0.50 | 91665 | 5.7 | 10.9 |
| Delaware | 0.40 | 81361 | 6.5 | 22.5 |
| District of Columbia | 1.20 | 108210 | 2.7 | 40.9 |
| Florida | 0.40 | 73311 | 10.7 | 14.9 |
| Georgia | 0.30 | 74632 | 11.4 | 30.8 |
| Idaho | 0.50 | 74942 | 8.9 | 0.8 |
| Illinois | 0.50 | 80306 | 6.2 | 13.3 |
| Indiana | 0.80 | 69477 | 6.9 | 9 |
| Iowa | 0.30 | 71433 | 5 | 4 |
| Kansas | 0.40 | 70333 | 8.4 | 5.3 |
| Kentucky | 1.30 | 61118 | 5.4 | 7.5 |
| Louisiana | 0.40 | 58229 | 6.9 | 30.3 |
| Maine | 0.60 | 73733 | 5.9 | 1.8 |
| Maryland | 0.80 | 98678 | 6.3 | 29.2 |
| Massachusetts | 0.60 | 99858 | 2.6 | 7 |
| Michigan | 0.70 | 69183 | 4.5 | 13.2 |
| Minnesota | 0.30 | 85086 | 4.2 | 7.2 |
| Mississippi | 0.40 | 54203 | 10.3 | 35.6 |
| Missouri | 0.60 | 68545 | 7.5 | 10.8 |
| Nevada | 0.60 | 76364 | 10.8 | 9.4 |
| New Jersey | 1.20 | 99781 | 7.2 | 12.7 |
| New Mexico | 1.60 | 62268 | 9.1 | 2 |
| New York | 0.60 | 82095 | 4.8 | 14.3 |
| North Carolina | 0.60 | 70804 | 9.2 | 20.1 |
| Ohio | 0.80 | 67769 | 6.1 | 11.9 |
| Oklahoma | 0.60 | 62138 | 11.4 | 6.8 |
| Oregon | 0.70 | 80160 | 5.5 | 2.1 |
| Pennsylvania | 0.70 | 73824 | 5.4 | 10.6 |
| Rhode Island | 1.40 | 84972 | 4.5 | 5.4 |
| South Carolina | 1.10 | 67804 | 9.1 | 24.4 |
| Tennessee | 1.20 | 67631 | 9.3 | 15.3 |
| Texas | 0.30 | 75780 | 16.4 | 12.3 |
| Utah | 0.60 | 93421 | 8 | 1.1 |
| Virginia | 0.80 | 89931 | 6.4 | 18.4 |
| Washington | 0.90 | 94605 | 6.3 | 4 |
| West Virginia | 0.90 | 55948 | 5.9 | 3.2 |
| Wisconsin | 0.40 | 74631 | 4.9 | 5.9 |

^a^United States Centers for Disease Control and Prevention (CDC). Naloxone dispensing rate maps. h[ttps://www.cdc.gov/overdose-prevention/data-research/facts-stats/naloxone-dispensing-rate-maps.html](https://www.cdc.gov/overdose-prevention/data-research/facts-stats/naloxone-dispensing-rate-maps.html);

^b^American Community Survey 2023 (United States Census Bureau)

**Table S5:** Correlation matrix for state-level annual percent change (APC) in polydrug OD mortality in 2024 versus 2023 with sociodemographic / public health variables. Variables are examined in a multiple linear regression (Table 2 in manuscript). Spearman r and uncorrected p-values are shown.

| **Variables** | **APC in 2024 versus 2023** | | **Naloxone prescriptions dispensed/100 persons (2023)** | | **Median household income**  **(2023)** | | **%Without health insurance coverage**  **(2023)** | | **%Population identifying as African-American or Black (2023)** | |
| --- | --- | --- | --- | --- | --- | --- | --- | --- | --- | --- |
|  | r value | p-value | r value | p-value | r value | p-value | r value | p-value | r value | p-value |
| **APC in 2024 versus 2023** | N/A | |  | | | | | | | |
| **Naloxone prescriptions dispensed / 100 persons (2023)** | -0.124 | NS | N/A | |  | | | | | |
| **Median household income**  **(2023)** | 0.296 | 0.0057 | -0.001 | NS | N/A | |  | | | |
| **%Without health insurance coverage**  **(2023)** | 0.305 | 0.0043 | -0.181 | 0.096 | -0.320 | 0.003 | N/A | |  | |
| **%Population identifying as**  **African-American or Black**  **(2023)** | -0.386 | 0.00025 | -0.069 | NS | -0.142 | NS | 0.134 | NS | N/A | |
| **Polydrug OD mortality rate/100,000**  **(2023)** | -0.238 | 0.027 | 0.253 | 0.019 | 0.019 | NS | -0.151 | NS | -0.001 | NS |

**Table S6:**  Age stratification of polydrug synthetic opioid *and* stimulant OD crude mortality rates/100,000 population: National data stratified in 10-year age groups and optimal models from joinpoint regression. *Indicates that modeled Annual Percent Change is significantly different from 0.

| **Year** | **Sex** | **Age** | **Deaths** | **Population** | **Observed Crude Rate** | **SE** | **Modeled Crude Rate** | **Joinpoint Location** | **Modeled Annual Percent Change** |
| --- | --- | --- | --- | --- | --- | --- | --- | --- | --- |
| 2018 | Female | 15-24 | 284 | 20994345 | 1.35 | 0.08 | 1.17 |  | 40.9426* |
| 2019 | Female | 15-24 | 295 | 20877151 | 1.41 | 0.08 | 1.65 |  | 40.9426* |
| 2020 | Female | 15-24 | 496 | 20828241 | 2.38 | 0.11 | 2.33 |  | 40.9426* |
| 2021 | Female | 15-24 | 660 | 21092449 | 3.13 | 0.12 | 3.28 | Joinpoint 1 |  |
| 2022 | Female | 15-24 | 626 | 21657540 | 2.89 | 0.12 | 2.86 |  | -12.9303* |
| 2023 | Female | 15-24 | 640 | 21463206 | 2.98 | 0.12 | 2.49 |  | -12.9303* |
| 2024 | Female | 15-24 | 358 | 21463206 | 1.67 | 0.09 | 2.16 |  | -12.9303* |
| 2018 | Male | 15-24 | 559 | 21976455 | 2.54 | 0.11 | 2.3 |  | 52.0736* |
| 2019 | Male | 15-24 | 676 | 21810359 | 3.1 | 0.12 | 3.5 |  | 52.0736* |
| 2020 | Male | 15-24 | 1103 | 21727443 | 5.08 | 0.15 | 5.32 | Joinpoint 1 |  |
| 2021 | Male | 15-24 | 1285 | 21996214 | 5.84 | 0.16 | 5.3 |  | -0.43 |
| 2022 | Male | 15-24 | 1214 | 22684031 | 5.35 | 0.15 | 5.28 |  | -0.43 |
| 2023 | Male | 15-24 | 1112 | 22423446 | 4.96 | 0.15 | 5.25 | Joinpoint 2 |  |
| 2024 | Male | 15-24 | 625 | 22423446 | 2.79 | 0.11 | 2.75 |  | -47.5760* |
| **Year** | **Sex** | **Age** | **Deaths** | **Population** | **Observed Crude Rate** | **SE** | **Modeled Crude Rate** | **Joinpoint Location** | **Modeled Annual Percent Change** |
| 2018 | Female | 25-34 | 1124 | 22487065 | 5 | 0.15 | 4.88 |  | 28.8403* |
| 2019 | Female | 25-34 | 1302 | 22581141 | 5.77 | 0.16 | 6.29 |  | 28.8403* |
| 2020 | Female | 25-34 | 1912 | 22625267 | 8.45 | 0.19 | 8.1 |  | 28.8403* |
| 2021 | Female | 25-34 | 2538 | 22441743 | 11.31 | 0.22 | 10.44 |  | 28.8403* |
| 2022 | Female | 25-34 | 2656 | 22311738 | 11.9 | 0.23 | 13.45 | Joinpoint 1 |  |
| 2023 | Female | 25-34 | 2571 | 22483329 | 11.44 | 0.23 | 10.44 |  | -22.3207* |
| 2024 | Female | 25-34 | 1691 | 22483329 | 7.52 | 0.18 | 8.11 |  | -22.3207* |
| 2018 | Male | 25-34 | 2440 | 23210709 | 10.51 | 0.21 | 10.57 |  | 32.5267* |
| 2019 | Male | 25-34 | 2912 | 23359180 | 12.47 | 0.23 | 14.01 |  | 32.5267* |
| 2020 | Male | 25-34 | 4688 | 23444379 | 20 | 0.29 | 18.57 |  | 32.5267* |
| 2021 | Male | 25-34 | 6153 | 23053362 | 26.69 | 0.34 | 24.61 |  | 32.5267* |
| 2022 | Male | 25-34 | 6563 | 23189562 | 28.3 | 0.35 | 32.62 | Joinpoint 1 |  |
| 2023 | Male | 25-34 | 6393 | 23059187 | 27.72 | 0.35 | 24.23 |  | -25.7097* |
| 2024 | Male | 25-34 | 3721 | 23059187 | 16.14 | 0.26 | 18 |  | -25.7097* |
| **Year** | **Sex** | **Age** | **Deaths** | **Population** | **Observed Crude Rate** | **SE** | **Modeled Crude Rate** | **Joinpoint Location** | **Modeled Annual Percent Change** |
| 2018 | Female | 35-44 | 939 | 20690288 | 4.54 | 0.15 | 4.35 |  | 41.1577* |
| 2019 | Female | 35-44 | 1124 | 20867064 | 5.39 | 0.16 | 6.15 |  | 41.1577* |
| 2020 | Female | 35-44 | 1943 | 21090324 | 9.21 | 0.21 | 8.68 |  | 41.1577* |
| 2021 | Female | 35-44 | 2899 | 21546241 | 13.45 | 0.25 | 12.25 |  | 41.1577* |
| 2022 | Female | 35-44 | 3255 | 21575176 | 15.09 | 0.26 | 17.29 | Joinpoint 1 |  |
| 2023 | Female | 35-44 | 3385 | 22028457 | 15.37 | 0.26 | 13.87 |  | -19.7755* |
| 2024 | Female | 35-44 | 2242 | 22028457 | 10.18 | 0.21 | 11.12 |  | -19.7755* |
| 2018 | Male | 35-44 | 2269 | 20587600 | 11.02 | 0.23 | 10.1 |  | 58.1189* |
| 2019 | Male | 35-44 | 2983 | 20792080 | 14.35 | 0.26 | 15.98 |  | 58.1189* |
| 2020 | Male | 35-44 | 4909 | 21045868 | 23.33 | 0.33 | 25.26 | Joinpoint 1 |  |
| 2021 | Male | 35-44 | 7172 | 21857613 | 32.81 | 0.39 | 29.96 |  | 18.5906* |
| 2022 | Male | 35-44 | 8229 | 22120189 | 37.2 | 0.41 | 35.53 |  | 18.5906* |
| 2023 | Male | 35-44 | 8890 | 22362236 | 39.75 | 0.42 | 42.13 | Joinpoint 2 |  |
| 2024 | Male | 35-44 | 5567 | 22362236 | 24.89 | 0.33 | 24.7 |  | -41.3738* |
| **Year** | **Sex** | **Age** | **Deaths** | **Population** | **Observed Crude Rate** | **SE** | **Modeled Crude Rate** | **Joinpoint Location** | **Modeled Annual Percent Change** |
| 2018 | Female | 45-54 | 656 | 21090497 | 3.11 | 0.12 | 3.18 |  | 41.7206* |
| 2019 | Female | 45-54 | 846 | 20702936 | 4.09 | 0.14 | 4.51 |  | 41.7206* |
| 2020 | Female | 45-54 | 1367 | 20441441 | 6.69 | 0.18 | 6.39 |  | 41.7206* |
| 2021 | Female | 45-54 | 2044 | 20376477 | 10.03 | 0.22 | 9.05 |  | 41.7206* |
| 2022 | Female | 45-54 | 2232 | 20152015 | 11.08 | 0.23 | 12.83 | Joinpoint 1 |  |
| 2023 | Female | 45-54 | 2395 | 20306672 | 11.79 | 0.24 | 10.34 |  | -19.4019 |
| 2024 | Female | 45-54 | 1531 | 20306672 | 7.54 | 0.19 | 8.34 |  | -19.4019 |
| 2018 | Male | 45-54 | 1685 | 20541202 | 8.2 | 0.2 | 7.55 |  | 61.5054* |
| 2019 | Male | 45-54 | 2227 | 20171966 | 11.04 | 0.23 | 12.2 |  | 61.5054* |
| 2020 | Male | 45-54 | 3605 | 19924692 | 18.09 | 0.3 | 19.7 | Joinpoint 1 |  |
| 2021 | Male | 45-54 | 5278 | 20311959 | 25.98 | 0.36 | 23.79 |  | 20.7610* |
| 2022 | Male | 45-54 | 6169 | 20279630 | 30.42 | 0.39 | 28.73 |  | 20.7610* |
| 2023 | Male | 45-54 | 6569 | 20187109 | 32.54 | 0.4 | 34.7 | Joinpoint 2 |  |
| 2024 | Male | 45-54 | 4152 | 20187109 | 20.57 | 0.32 | 20.42 |  | -41.1586* |
| **Year** | **Sex** | **Age** | **Deaths** | **Population** | **Observed Crude Rate** | **SE** | **Modeled Crude Rate** | **Joinpoint Location** | **Modeled Annual Percent Change** |
| 2018 | Female | 55-64 | 366 | 21873773 | 1.67 | 0.09 | 1.57 |  | 54.6395* |
| 2019 | Female | 55-64 | 488 | 21949318 | 2.22 | 0.1 | 2.43 |  | 54.6395* |
| 2020 | Female | 55-64 | 814 | 21914243 | 3.71 | 0.13 | 3.77 |  | 54.6395* |
| 2021 | Female | 55-64 | 1467 | 21839746 | 6.72 | 0.18 | 5.82 |  | 54.6395* |
| 2022 | Female | 55-64 | 1633 | 21413534 | 7.63 | 0.19 | 9 | Joinpoint 1 |  |
| 2023 | Female | 55-64 | 1801 | 21350566 | 8.44 | 0.2 | 7.46 |  | -17.1571 |
| 2024 | Female | 55-64 | 1169 | 21350566 | 5.48 | 0.16 | 6.18 |  | -17.1571 |
| 2018 | Male | 55-64 | 970 | 20398863 | 4.76 | 0.15 | 4.44 |  | 71.9243* |
| 2019 | Male | 55-64 | 1442 | 20499219 | 7.03 | 0.19 | 7.63 |  | 71.9243* |
| 2020 | Male | 55-64 | 2387 | 20489434 | 11.65 | 0.24 | 13.11 | Joinpoint 1 |  |
| 2021 | Male | 55-64 | 3933 | 20963318 | 18.76 | 0.3 | 17.3 |  | 31.8911* |
| 2022 | Male | 55-64 | 5060 | 20671903 | 24.48 | 0.34 | 22.81 |  | 31.8911* |
| 2023 | Male | 55-64 | 5769 | 20503845 | 28.14 | 0.37 | 30.09 | Joinpoint 2 |  |
| 2024 | Male | 55-64 | 3395 | 20503845 | 16.56 | 0.28 | 16.43 |  | -45.3853* |
| **Year** | **Sex** | **Age** | **Deaths** | **Population** | **Observed Crude Rate** | **SE** | **Modeled Crude Rate** | **Joinpoint Location** | **Modeled Annual Percent Change** |
| 2018 | Female | 65-74 | 35 | 16246231 | 0.22 | 0.04 | 0.24 |  | 55.6232* |
| 2019 | Female | 65-74 | 61 | 16783854 | 0.36 | 0.05 | 0.38 |  | 55.6232* |
| 2020 | Female | 65-74 | 93 | 17365858 | 0.54 | 0.06 | 0.59 |  | 55.6232* |
| 2021 | Female | 65-74 | 174 | 17797036 | 0.98 | 0.07 | 0.91 |  | 55.6232* |
| 2022 | Female | 65-74 | 273 | 17877767 | 1.53 | 0.09 | 1.42 |  | 55.6232* |
| 2023 | Female | 65-74 | 372 | 18350204 | 2.03 | 0.11 | 2.21 | Joinpoint 1 |  |
| 2024 | Female | 65-74 | 278 | 18350204 | 1.51 | 0.09 | 1.47 |  | -33.3791 |
| 2018 | Male | 65-74 | 174 | 14246085 | 1.22 | 0.09 | 1.29 |  | 50.7384* |
| 2019 | Male | 65-74 | 253 | 14699579 | 1.72 | 0.11 | 1.94 |  | 50.7384* |
| 2020 | Male | 65-74 | 433 | 15183540 | 2.85 | 0.14 | 2.92 |  | 50.7384* |
| 2021 | Male | 65-74 | 767 | 15869086 | 4.83 | 0.17 | 4.4 |  | 50.7384* |
| 2022 | Male | 65-74 | 1108 | 15910672 | 6.96 | 0.21 | 6.64 |  | 50.7384* |
| 2023 | Male | 65-74 | 1546 | 16335080 | 9.46 | 0.24 | 10 | Joinpoint 1 |  |
| 2024 | Male | 65-74 | 1092 | 16335080 | 6.68 | 0.2 | 6.64 |  | -33.6474 |
